# The parallel auditory brainstem response paradigm provides accurate and fast hearing thresholds in a clinic-like setting

**DOI:** 10.1101/2025.11.26.25341073

**Authors:** Melissa J Polonenko, Ross K Maddox

**Affiliations:** Department of Speech-Language-Hearing Sciences, University of Minnesota; Kresge Hearing Research Institute, Dept. of Otolaryngology—Head and Neck Surgery, University of Michigan

## Abstract

**Objectives:** The auditory brainstem response (ABR) is an essential tool in screening for and diagnosing infant hearing loss, and its results drive decisions regarding interventions and hearing habilitation with impacts extending far into a child’s future. Despite the traditional ABR exam’s usefulness, there is an identified need to develop faster, more informative exams. The parallel ABR (pABR) measures responses to all frequencies of interest in both ears all at once, rather than the traditional series of single-frequency measurements in one ear at a time, greatly speeding the diagnostic exam. The pABR has been shown to be effective at quickly measuring frequency-specific responses in adults with normal hearing, but it has not yet been tested in people with hearing loss. The goal of this study was to determine the accuracy and speed of the pABR for estimating hearing thresholds in a clinic-like setting.

**Design:** Seventy adults with widely varying sensorineural hearing loss configurations were recruited to participate in this study. We measured thresholds at octave frequencies in two ways: the behavioral audiogram, serving as the ground truth, and using the pABR with a custom-designed interactive user interface. Accuracy was determined through threshold correlation coefficients as well as absolute error in decibels. Acquisition time was assessed as the time from measurement start to determination of the final threshold. To determine the pABR’s speed advantages, a subset of participants was invited back and their thresholds estimated a third time, using a commercially available clinical system to serially measure ABR waveforms. Speedup was assessed in terms of the raw difference in acquisition time in minutes and as the ratio between measurement times made with the two ABR paradigms.

**Results:** Thresholds estimated with pABR highly correlated with the behavioral audiogram ground truth. The correlation was 0.90 (0.88–0.92, 95% confidence interval) across all ears and frequencies. 79% of pABR thresholds were within one 10-dB step-size of the behavioral threshold. The pABR was faster in all ten participants where traditional serial ABR was also recorded, with a mean recording time of 28 minutes to estimate ten pABR thresholds (500–8000 Hz in each ear) versus 70 minutes to estimate eight serial thresholds (500–4000 Hz in each ear), or a mean reduction of 42 minutes. The median speedup ratio was 2.5×.

**Conclusions:** The pABR provides accurate threshold estimates with greatly reduced measurement time compared to traditional methods. Given these results and other advantages related to its design, the pABR holds promise as a clinical tool that can be deployed to commercial systems in the near future.

## Introduction

The diagnostic auditory brainstem response (ABR) is an essential step in the process of early hearing loss detection and intervention. It uses the presence or absence of an evoked potential in response to transient acoustic stimuli at varying frequencies and levels to estimate hearing thresholds in infants and other people who cannot perform behavioral audiometry (Hood, 1998). Under ideal recording conditions, thresholds estimated with ABR strongly predict gold standard behavioral thresholds (Stapells and Oates, 1997; McCreery et al., 2015). Practical conditions, however, are often far from ideal. The ABR is small. High levels of noise in a still patient necessitate many thousand stimulus repetitions before a clear response emerges. A recording in which the patient is moving or fidgeting may not be usable at all. To reduce these movement artifacts, the exam is typically done in infants while they sleep (Janssen et al., 2010). This requirement, along with the fact that many measurements need to be made at different ear-frequency-level combinations, is the source of the major practical constraint on the diagnostic ABR: time.

The parallel ABR (pABR) is a paradigm that allows the simultaneous collection of responses to stimuli at all frequencies in both ears in parallel (Polonenko and Maddox, 2019). By measuring many responses at once, the set of all responses can be collected much faster when used in the lab (Polonenko and Maddox, 2019; Suphinnapong et al., 2025). This speed is the pABR’s primary advantage, but it has others. The pABR is flexible: it is effective with widely varying stimulus presentation rates (Polonenko and Maddox, 2021), the analysis window can be extended to include the middle latency response (MLR; Polonenko and Maddox, 2021), and while the published work so far has used tonebursts, other stimuli such as chirps could also be employed. A pair of studies has also shown through modeling and electroencephalography (EEG) that the responses obtained from the pABR are more place-specific than those acquired through serial presentation (Stoll and Maddox, 2023, 2024).

The primary goal for the pABR is its development as a clinical tool. Important questions remain to that end and answering them is the goal of this paper. The first question is of accuracy: does the pABR accurately predict gold standard behavioral audiometric thresholds in people with widely varying hearing loss configurations? Traditional serial ABR is known to be accurate, but it needs to be shown that the differences introduced by the pABR (simultaneous response measurement and randomized stimulus timing) do not impact this accuracy. The second question is of speed: how much faster is the pABR than the standard protocol? Since traditional ABR is accurate, the pABR is only worth translating if it offers a significant speed advantage.

For this study we recruited 70 adult human participants with widely varying sensorineural hearing loss. We estimated their hearing thresholds from 500 to 8000 Hz in two ways: first the pABR and then behavioral pure tone audiometry. The pABR thresholds, which we measured using a custom interactive clinical interface, were strongly correlated with behavioral thresholds, confirming that the pABR indeed provides accurate threshold estimates across different severities and configurations of hearing loss. We also recruited a subset of 10 participants to return to the lab for a standard ABR exam using commercially available equipment to compare the speed of threshold estimates. We found that the pABR was faster than the standard protocol in every participant and was on average 2.5 times faster. This speed advantage came despite measuring ten thresholds with the pABR (500–8000 Hz in each ear) but only eight with the standard protocol (500–4000 Hz in each ear).

This study adds important findings to the existing literature in simultaneous ABR measurement. It establishes that the pABR can be measured interactively in a clinic-like setting in people with hearing loss, it shows that those measurements are highly accurate, and it demonstrates that those measurements can be made much more quickly than using traditional ABR techniques. These findings pave the way for the pABR’s transition from the lab to the clinic.

## Methods

### Participant recruitment

Seventy participants were recruited. Four were recruited at the University of Rochester, under a protocol approved by the University of Rochester Research Participants Review Board (STUDY00003866). The remaining 66 were recruited at the University of Minnesota under a single IRB protocol (SITE00000908) overseen by the University of Michigan IRBMED (HUM00252149) with the IRB of the University of Minnesota signing on. All participants gave written informed consent. All participants were compensated for their time.

Participants were recruited to have widely varying hearing loss configurations. Participants’ ages ranged from 18 to 70 years, with a median (first, third quartiles) of 54 (39, 62) years. There were 41 females and 29 males. The limit of our equipment was 100 dB peSPL before audio clipping. Thus, to ensure at least some measurable pABR responses, participants were included if they had hearing thresholds <85 dB HL at most frequencies in both ears. There were two exceptions: we recruited two participants who had one deaf ear to ensure spurious thresholds were not logged in the better hearing ear with the simultaneous presentation paradigm.

Ten participants who participated in the main experiment returned to the lab so that their thresholds could be estimated with a commercial ABR system. These participants were chosen through an algorithm that computed the Euclidean distance between participants’ thresholds in the ten-dimensional audiogram space and found a subset that led to a high total inter-participant distance. The effect was that these ten participants had mutually dissimilar audiograms. This analysis was performed at the University of Michigan and the researchers collecting data at the University of Minnesota were blind to the thresholds of those returning participants.

### pABR stimuli and response collection

The pABR was measured using previously described methods (Polonenko and Maddox, 2019). In brief, pABR stimuli were constructed from five-cycle Blackman-windowed tonebursts centered at octave frequencies from 500 Hz to 8000 Hz. For each frequency, a toneburst train was created by placing tonebursts randomly within a 1 s epoch. This was repeated for all other frequencies with independent random processes controlling the timing. All toneburst trains were summed, and the process repeated with new random processes for the other ear, comprising a stimulus epoch. Figure 1 provides an overview of this process. Because of the independent timing, the average ABR waveform to each toneburst train could be computed from the same EEG data free from interference. Responses were computed as the cross-correlation of each stimulus timing sequence divided by the number of stimuli in that trial (this is equivalent to standard epoching and averaging but more efficient to compute). The stimulus presentation rate was 40 stimuli / s for each frequency-ear, leading to an overall stimulus rate of 400 stimuli / s. This stimulus rate has been shown to efficiently provide responses to all frequencies (Polonenko and Maddox, 2021). By varying the level of these sets of toneburst trains, thresholds were determined for each frequency-ear combination. The levels of all stimuli were varied together—stimuli at different frequencies were not independently changed. This resulted in some loud stimuli for steeply sloping hearing loss configurations. Measures were taken to limit exposure time to the loud levels, such as limiting testing time and returning to the level to gather sufficient epochs to determine the (higher) thresholds for the remaining frequencies.

**Figure 1.**
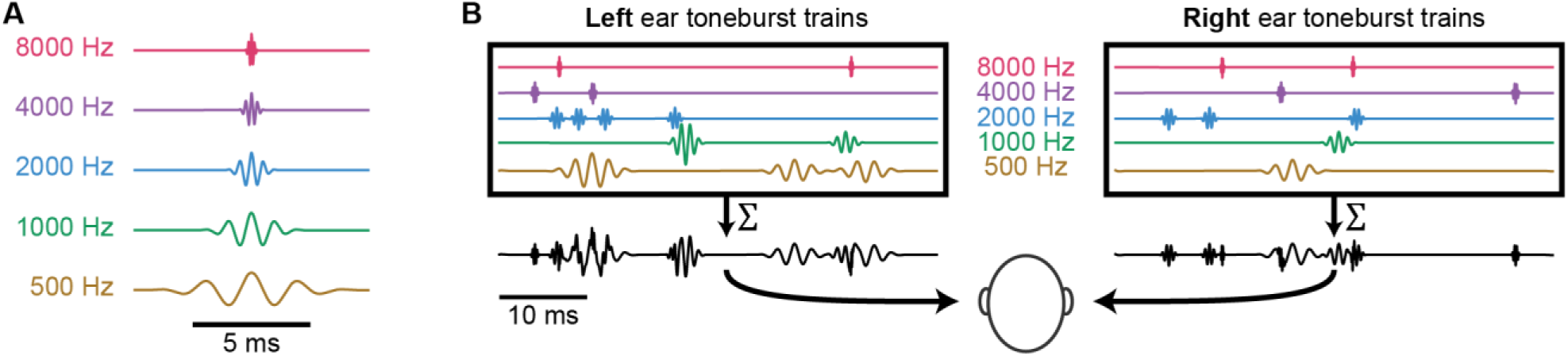
pABR stimulus construction. (**A**) Individual toneburst stimuli for each frequency. (**B**) Toneburst trains in each ear (colored lines) are summed to create a two-channel (left, right) stimulus epoch (black lines). From Polonenko & Maddox (2019), licensed CC BY-NC open access.

Stimuli were presented over insert earphones (Etymotic Research ER-2) plugged into a headphone amplifier (Schiit Audio Magni+), which received its signal from USB sound card (RME Digiface USB). Stimuli were presented at an audio sampling rate of 48,000 Hz.

EEG data were measured using a BrainVision ActiCHamp+ system with specialized ABR preamplifiers (EP-PreAmp). Data were streamed to the real-time interface using LabStreamingLayer (Kothe et al., 2025). Data were bandpass filtered between 30 and 1500 Hz using a causal first-order Butterworth filter. Bayesian averaging of responses was performed so that noisier data segments were down-weighted in the average (Polonenko and Maddox, 2019, 2021).

### Interface and search protocol

A custom interface was developed to allow real-time interactive testing. The interface allowed the experimenter to easily switch between levels and displayed all of the average waveforms collected so far. The experimenter could indicate a response by clicking on a waveform and the estimated hearing threshold levels in dB nHL were automatically updated and displayed using correction factors from Polonenko & Maddox (2021). The residual noise and signal-to-noise ratio (SNR) of each waveform was also estimated and displayed. The interface had several other options: data could be split into separate averages or viewing replications or separated compression / rarefaction polarity responses, and other aspects of the test (montage, filtering, analysis window time range, etc.) could also be changed on the fly. The interface was designed so that data collection never needed to be stopped (and thus no time wasted). This was aided by the fact that only the stimulus level (not the ear or the frequency) needed to be selected.

As in conventional ABR, wave V presence was the main indicator for thresholding. Unlike prior methods, the pABR also provides the MLR alongside the ABR. The MLR is a less consistent response than the ABR (Özdamar and Kraus, 1983; Kavanagh et al., 1989), making it inappropriate for threshold determination on its own, but we were able to use its presence or absence to resolve cases where wave V presence was ambiguous. We found this to be helpful especially at low frequencies (500 and 1000 Hz), which evoked broader responses. A response was marked present when a wave V was visible, or absent when nothing could be seen and a residual noise of ∼20 nV was obtained. As is often done in clinic, we ran levels at threshold, below threshold to confirm no response, and at least one level above threshold to confirm the response grew in size. In the case where the highest intensities were run to verify threshold, we limited the length of time we recorded until sufficient evidence was gained to confirm a response. To determine an absent response we recorded until the residual noise was ∼20 nV whenever possible.

The threshold search usually started at 70 dB peSPL, then we decided what levels to run next based on morphology across the frequencies. If there were no responses, or very small responses, we often ran a higher level to confirm an ambiguous response or to determine thresholds for the higher levels so as not to wake the person later. In this case, we then went down in level. If there were very robust responses, then we lowered the level to 40 dB peSPL to estimate whether the person had normal hearing (∼20 dB nHL), and then bracketed from there based on the size and morphology of the responses, as well as the participant’s rest state (i.e., if very quietly sleeping, we ran the lower levels to ensure we had low noise for the very quiet levels before increasing the level again so as to not wake the individual). We implemented a “warning” button for levels >80 dB peSPL, such that when selecting higher levels, we had to click the button to acknowledge that we were to limit the duration of continuous data collection at the higher levels. Because we could combine different runs of the same level in our displayed waveforms, we often alternated between a lower and higher level to give breaks from the higher intensities while still collecting sufficient data to make a judgment about whether a response was present or absent. In some cases, we only collected a few trials at the highest level if we were only determining if the response grew in amplitude and confirm the next level down was threshold.

### Regressions and correction factors

ABR thresholds require some form of conversion before they can be compared with behavioral thresholds. We first converted from dB peSPL to dB nHL using our previously established correction factors (Polonenko and Maddox, 2021). Correcting from dB nHL to dB estimated HL (eHL) can be done using slope-intercept regression (McCreery et al., 2015) or as additive correction factors (Gorga et al., 2006; Vander Werff et al., 2009; Stapells, 2011). The latter were computed as the mean dB HL − dB nHL difference. For the sake of completeness, we computed both types of corrections. The additive correction factors were similar enough across frequencies that we also computed a single additive correction factor for all frequencies. Thresholds that were higher than could be determined with our equipment were excluded from correction factor computation and accuracy analyses.

To compare the accuracy of the pABR in predicting behavioral thresholds we computed correlations between the thresholds as well as absolute errors. To make these comparisons we fit slope-intercept models for each frequency to convert the pABR thresholds to dB eHL as above, with the exception that we used a leave-one-participant-out approach. These unbiased corrections are used in Figure 2.

**Figure 2.**
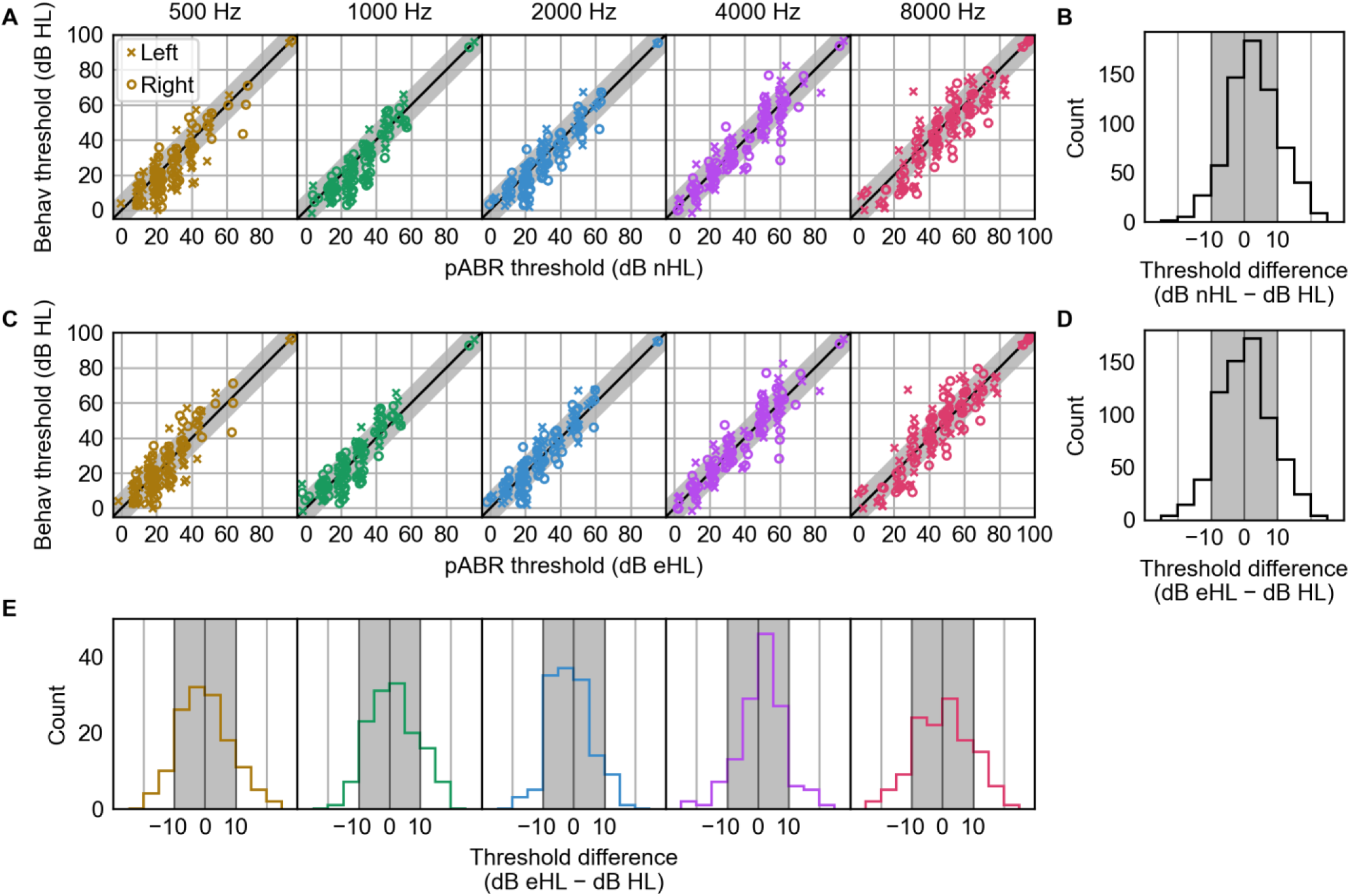
Comparison of pABR thresholds to behavioral audiogram thresholds. (**A**) Behavioral audiogram (in dB HL) versus pABR thresholds in dB nHL (i.e., before correction applied) for each frequency. Jitter applied for visualization (±2.5 dB). Shaded areas show differences within the step size of ±10 dB. Thresholds shown as 95 dB in cases where they could not be determined at the highest sitmulus level (these points are not included in panels B, D, or E). (**B**) Histogram of errors across all frequencies for thresholds expressed in dB nHL. (**C**) Comparison between behavioral thresholds and pABR thresholds in dB eHL (i.e., after correction using regression with a leave-one-out approach). Plotted as in A. (**D**) Errors for corrected thresholds across all frequencies. (**E**) Errors for corrected thresholds, separated by frequency.

## Results

### Accuracy

Overall, pABR threshold estimates agreed quite well with behavioral pure tone audiograms in dB HL. Figure 2 shows the comparisons for pABR thresholds in dB nHL (Figure 2A, B) and dB eHL (Figure 2C–E). We focus our accuracy analyses on thresholds in dB eHL, computed in a leave-one-out fashion as described above. Across all frequencies, the overall correlation between pABR and behavioral thresholds was 0.90 (95% confidence interval, 0.89–0.92), which is in line with traditional toneburst ABR (Stapells and Oates, 1997; Gorga et al., 2006; Ramos et al., 2013). Correlations varied slightly for individual frequencies, but were still high for all: 0.83 at 500 Hz, 0.87 at 1000 Hz, 0.91 at 2000 Hz, 0.92 at 4000 Hz, and 0.88 at 8000 Hz. Correlations were not different when pABR thresholds were expressed in dB nHL.

The error between pABR and behavioral thresholds was also low (Figure 2D). The median (first, third quartiles) for the difference (dB eHL – dB HL) in estimates was 0.2 dB (−5.1, 5.3 dB) using the leave-one-out regression approach. The median absolute error was 5.3 dB (2.5, 9.3 dB), and 79% of pABR thresholds were within 10 dB (one step size) of the behavioral threshold, and 98.5% were within two step sizes (20 dB). Per-frequency errors are shown in Figure 2E. The median (first, third quartiles) error was 1.5 dB (−6.4, 5.8 dB) at 500 Hz, 0.2 dB (−4.9, 5.5 dB) at 1000 Hz, −1.1 dB (−6.1, 3.8 dB) at 2000 Hz, 0.7 dB (−4.3, 5.5 dB) at 4000 Hz, and 0.2 (−5.5, 6.4 dB) at 8000 Hz. The median absolute error was 6.2 dB (2.6, 9.0 dB) at 500 Hz, 5.0 dB (3.6, 9.6 dB) at 1000 Hz, 4.0 dB (2.7, 7.3 dB) at 2000 Hz, 4.7 dB (1.1, 9.0 dB) at 4000 Hz, and 5.3 dB (2.5, 9.3 dB) at 8000 Hz.

So far, we have considered the threshold data in aggregate, but a diagnostic measure must be effective in individuals for it to be useful. Figure 3 overlays the pABR and behavioral threshold estimates for every participant in the study. It can be qualitatively appreciated from that figure that the pABR threshold estimates are accurate across the range of hearing loss severities. The configuration of hearing loss also does not seem to matter, with closely matching estimates for steeply sloping loss (participants 14, 30, 53), low frequency loss (3, 6, 52, 63), notched loss (20, 48), asymmetric loss (3, 6, 11, 14, 15, 23, 24, 39, 63), or even single-sided deafness (7). For the participant with single-sided deafness, there were no responses from the deaf ear except at the highest levels, but these responses were not taken as threshold, rather considered crossover given the ∼70 dB difference in levels between ears. The matching results across configurations of hearing loss suggest that the inherent “masking” of the pABR stimuli works when presenting in both ears to a large extent. For low-frequency loss where there would be upward spread of masking from the stimuli, the estimates of threshold still correlated, consistent with work that shows the parallel stimuli improve place specificity at higher levels (Stoll and Maddox, 2023, 2024).

**Figure 3.**
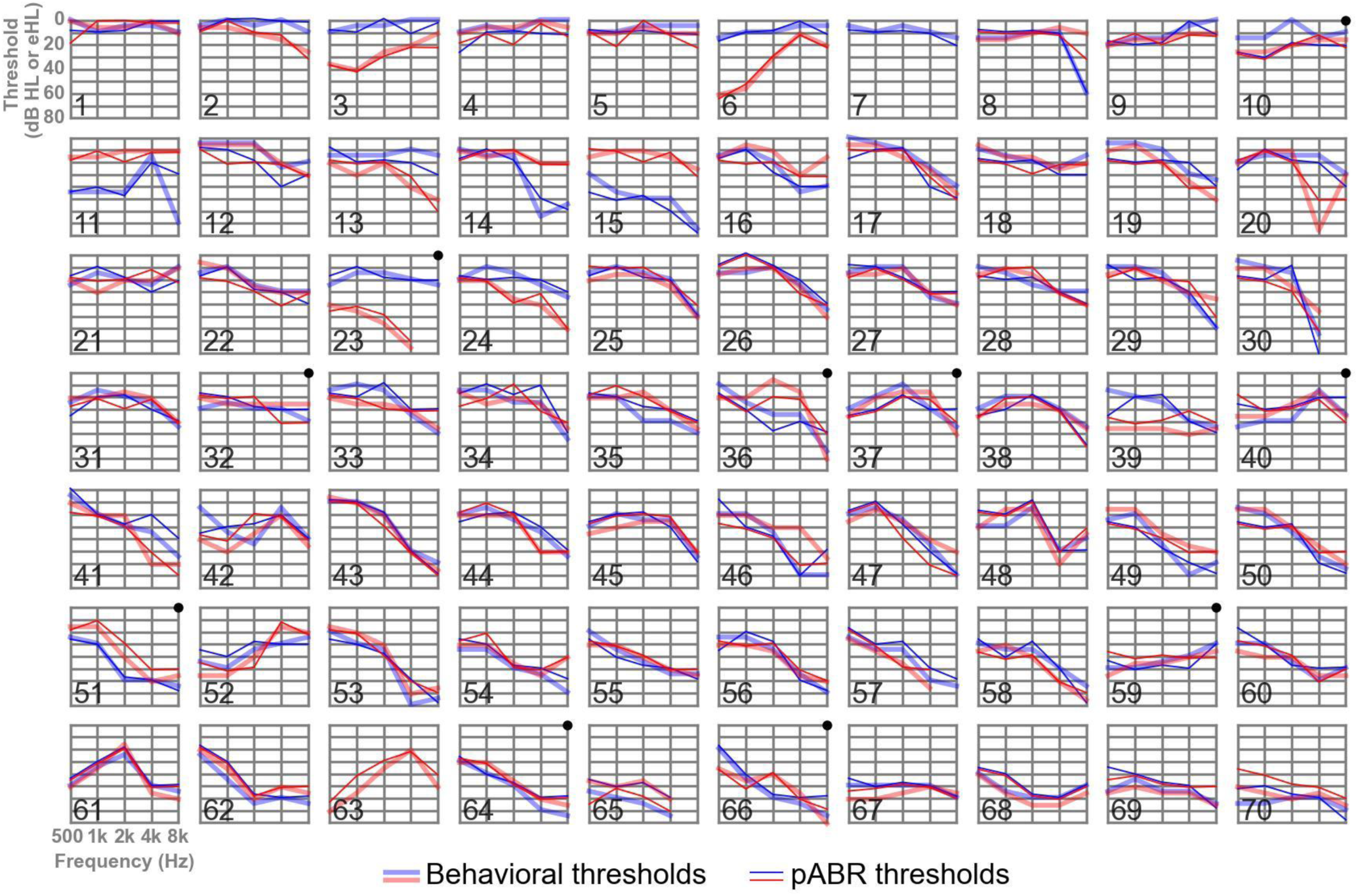
Audiograms and pABR thresholds for all study participants, arranged roughly in order of increasing severity of the better ear. Behavioral audiograms (in dB HL) are shown in thick, transparent lines. Thresholds estimated from pABR (in dB eHL) are shown in thin, solid lines. Left and right ears are shown in blue and red, respectively. Ten participants who returned to the lab for testing with the commercial system are indicated by a black dot in the upper right corner. Absent datapoints indicate where a response could not be obtained for either the pABR or the audiogram at the highest level.

#### Correction factors

Figure 2A shows that the dB nHL thresholds are close to the unity line with the behavioral thresholds in dB HL over the range of hearing loss severities, and Figure 2B shows that the majority of nHL thresholds are within the 10-dB step size. Still, to get the most accurate threshold predictions, correction factors are needed. Corrections for converting thresholds in dB nHL to eHL can be done in multiple ways, depending on the level of accuracy required. From most detailed to least, they are a first-order slope-intercept fit for each frequency, an additive correction factor for each frequency, and a single additive correction factor for all frequencies. Figure 2A suggests that corrections from dB nHL to dB eHL should be small. This is borne out in the numbers.

The slope-intercept equations are

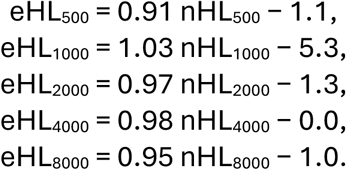

The per-frequency correction factors are

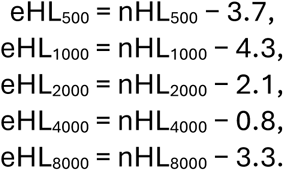

The equation for a single correction factor across all frequencies is

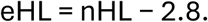

All three of these models explain a high degree of variance, at 99.98%, 99.72%, and 99.25%, respectively. Per-frequency additive corrections perform nearly as well as the slope-intercept fits because the slopes are all close to unity. Likewise, the single correction factor performs nearly as well as the per-frequency correction factors because the latter do not vary substantially across frequency.

### Speed

Having verified the pABR’s accuracy, its speed can now be considered. The time to collect a full set of ten thresholds varied widely (as it would in the clinic). The median (quartiles) time was 32 minutes (26, 37 minutes). Figure 4A shows a histogram of the time for all participants. The typical range was between 15 and 50 minutes, with a large mode in the low thirties and a median of 32 minutes. These times represent the true start-to-finish exam time, leading to a few participants taking nearly an hour—this typically occurred when signals were noisy to begin with and collection had to be stopped, adjustments made (e.g. to electrodes, determining source of noise), and measurement restarted.

**Figure 4.**
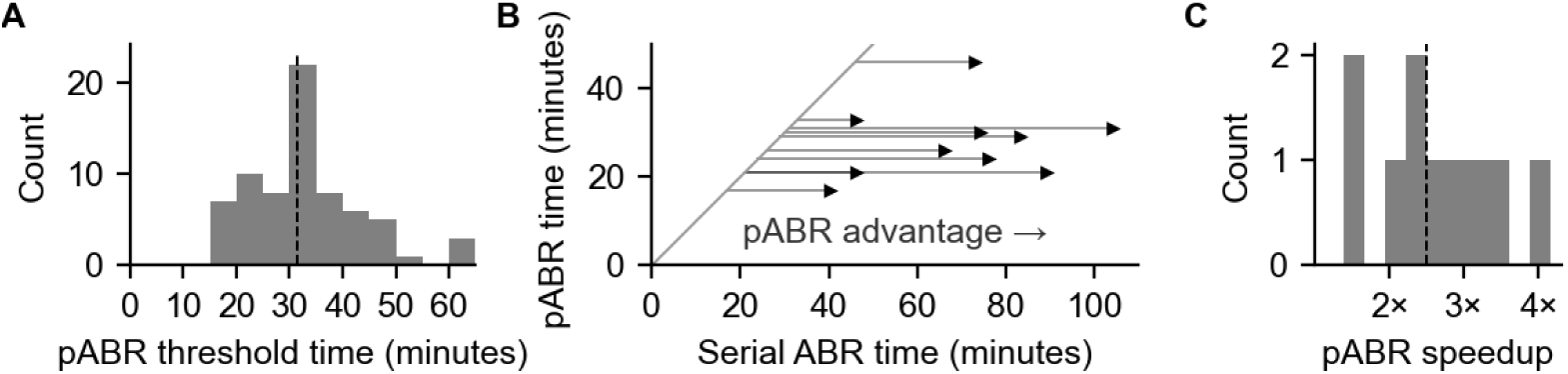
Recording times for pABR and comparison to serial ABR. (**A**) Distribution of time to estimate all thresholds for all 70 participants. Dashed line indicates median of 32 minutes. (**B**) pABR versus serial ABR threshold estimation times in ten adults with hearing loss who were recruited for the follow-up comparison experiment. All points are right of the diagonal, indicating pABR advantage. Length of horizontal line indicates improvement in minutes. (**C**) Distribution of pABR speed advantages versus serial. Dashed line indicates 2.5× median.

A subset of ten participants was recruited back to the lab so that their thresholds could also be estimated with a commercially available system (Interacoustics Eclipse) and the times compared with pABR directly. Parameters were matched as closely as possible and the threshold search procedure, including stopping criteria, were the same. While the pABR provided thresholds from 500 to 8000 Hz (ten thresholds per participant), the commercial system provided thresholds limited to 500 to 4000 Hz (eight thresholds). In those ten participants, the median time to estimate pABR thresholds was 28 minutes, compared to 74 minutes for the commercial system. The pABR was faster than the commercial system for all ten participants, with a median difference of 42 minutes (Figure 4B). The pABR was 1.4 to 4.2 times faster than the commercial system, with a median speedup of 2.5× (Figure 4C).

The times discussed so far have been to fully estimate all thresholds. In clinical use it is common to stop the exam as soon as normal hearing is confirmed. There were five participants whose hearing was determined to be normal at all frequencies in both ears by the pABR. In those participants, that assessment was made in a median time of 10 minutes. Times this short do not appear in the histogram of Figure 4A because we continued testing those participants so that thresholds could be determined in detail down to ∼0 dB nHL (20 dB peSPL), rather than stopping when thresholds were estimated to be normal (defined as 40 dB peSPL, or ∼20 dB nHL).

## Discussion

### Using the pABR for clinical threshold estimation

This study’s primary finding is that the pABR provides accurate hearing threshold estimates in substantially shorter times than conventional ABR for a wide range of hearing loss configurations. The correlation between pABR and behavioral thresholds was 0.90, in line with what has been found in prior studies of the conventional ABR, and pABR threshold estimation was 2.5× faster than thresholds obtained with the commercial system. These results support a compelling case for translation to the clinic. Had the pABR been accurate but only matched the conventional ABR in speed, there would be little reason to replace existing methods. Had it been inaccurate, it would not be worth pursuing, no matter how fast.

#### Accuracy

ABR threshold estimation at low frequencies can be difficult due to smaller, broader, and less defined waveforms (Stapells and Oates, 1997; Gorga et al., 2006). Prior work with the pABR has shown that low frequency responses, especially 500 Hz, are smaller still than serial ABR waveforms (Polonenko and Maddox, 2019). This size difference appears to stem from masking of those responses’ basal spread by the higher frequency stimuli being presented at the same time (Stoll and Maddox, 2023, 2024). We believe this improved place specificity has value, but it was a concern before completing this study that the smaller responses would mean thresholds at low frequencies were not sufficiently accurate, or that they would lead to exam times being too long. Here we saw that they were accurate (correlations of 0.83 and 0.87 for 500 and 1000 Hz, respectively) and it was still much faster to collect thresholds using pABR than serial. The presence of wave V is the key indicator when estimating thresholds, but the pABR allows additional information to be used. Unlike conventional ABR, where periodic stimulus presentation limits the analysis window to the inverse of the presentation rate, the pABR analysis window can be arbitrarily extended in either direction. We observed several instances where wave V was ambiguous but was followed by a clear MLR and made the choice to mark those responses as present and near/above threshold. This is a unique advantage of the pABR and helps to mitigate the small responses to low-frequency stimuli.

Bilaterally presenting the sound did not impact the pABR’s ability to distinguish asymmetric hearing losses. The amount of inherent masking was sufficient with the ER-2 insert earphones. In 10 cases with asymmetric thresholds, the pABR estimated the asymmetric loss (Figure 3). Furthermore, thresholds were not detected for the deaf ear in the two cases with confirmed deafness in one ear. Only for the one participant with a deaf ear and normal hearing in the other ear did very small waveforms appear at 100 dB peSPL for some of the frequencies, which were not determined to be threshold due suspicion of cross-over and the very low SNR, and thus not noted as threshold (participant 7 in Figure 3). While it is possible that bilateral stimulation led to some degree of contralateral suppression of responses (we did not directly test this), whatever suppression occurred neither prevented the pABR from being accurate nor dampened its sizeable speed advantage over serial presentation.

#### Speed

A chart review study by Jannsen et al. (2010) quantified the recording time available in infant patients as well as the number of thresholds obtained during that time. In naturally sleeping infants, a mean of 49 minutes were available, with 80% sleeping for at least 24 minutes. In those infants a mean of 6.2 thresholds were obtained, with at least 4.0 thresholds obtained from 80%. In the present study, the median pABR recording time across all participants was 32 minutes, during which 10 thresholds were obtained. The comparison between Jannsen et al. and our study is informative, but imperfect due to differences in the studies, most importantly the age of the participants. We made a direct comparison of pABR versus serial recording times in 10 of our adult participants, finding that the pABR was universally faster by a wide margin.

Based on the above, the pABR shows promise for providing shorter exams. Alternatively, clinicians may find that they still use the recording time available but acquire more measurements as a result, offering the possibility of better hearing aid fits when indicated. Either way, an important next step will be to validate the pABR in infants with hearing loss.

#### Exams that end before all thresholds are collected

In this study we collected all thresholds from all participants. In a clinical exam this is often the goal, but there are two main practical reasons why an exam might end early, the first voluntary and the second involuntary.

The first reason an exam may be stopped early is when normal thresholds can be confirmed by the audiologist. Though it will vary by hospital, this is the outcome in approximately half of infants (Janssen et al., 2010). In the case of normal thresholds (generally considered 20–25 dB HL or better) there is no clinical intervention indicated. It is thus not necessary to estimate thresholds beyond that point in most cases, and the patient can be sent home. Six participants in this study were determined to have normal hearing with the pABR. The median time to make that assessment was only 10 minutes, which corresponds to 1 minute per threshold. This is faster than could be done with any conventional system.

The second, involuntary reason why a session might be cut short when testing infants is if the patient wakes early. In serial ABR, the clinician would be left with whatever thresholds they measured up to that point and no information about other frequency-ear combinations. A truncated pABR session, on the other hand, provides at least some information about all thresholds. Sessions start at 50 dB eHL (70 dB peSPL) and move to 20 dB if all responses are obtained. If a session stops there, all ten thresholds are bracketed to ≤20 (normal), 30–50 dB (mild-moderate), or >50 (moderate-severe). Figure 5 shows two examples using data from this study, where data from only the first two levels tested were used, simulating early waking. One participant (#5 in Figure 3) is shown to have normal hearing, the other (#17 in Figure 3) is shown to have bilateral mild-moderate loss at 4000 and 8000 Hz. The morphology and robustness of the waveforms could provide further information about where thresholds are likely to be. Thus, while the pABR is still susceptible to early waking, more (and arguably more useful) information is obtained in a truncated pABR session than a serial ABR session of the same length.

**Figure 5.**
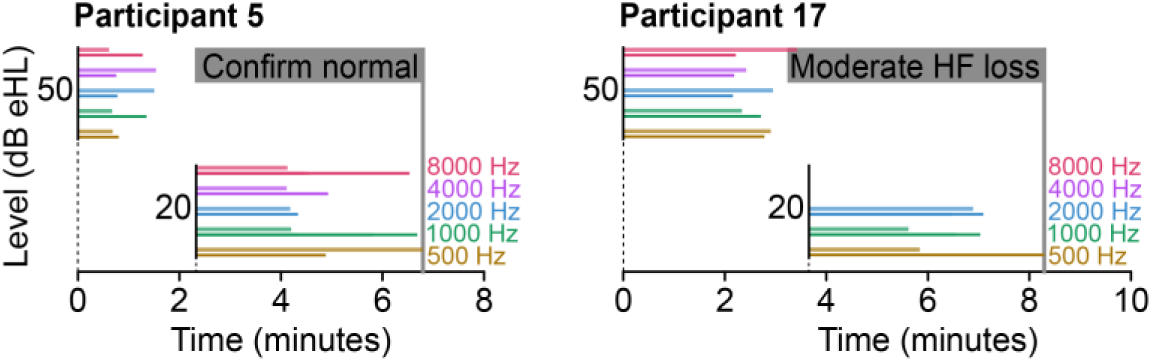
Two example pABR sessions truncated to the first two levels. Each colored line shows the time to indicate a reponse for that frequency-ear combination. Participant 5: normal thresholds confirmed in under 7 minutes. Participant 17: four of six high-frequency thresholds are normal in each ear, high-frequency thresholds are bracketed at 30–50 dB eHL in under 9 minutes.

### Comparison with the auditory steady-state response

The auditory steady-state response (ASSR) is an evoked response that is phase-locked to a periodic stimulus and can be measured with most commercial ABR systems. The stimulus is a tonal carrier at the test frequency (e.g., 500 Hz) whose amplitude is modulated in the 80–100 Hz range to create the subcortical steady-state response (Korczak et al., 2012). As with the toneburst ABR, correlations between ASSR and behavioral thresholds can be around 0.9 if a range of thresholds are considered (Luts et al., 2006). The multiple ASSR, like the pABR, can test more than one frequency and ear at a time. It does so by “tagging” tones with different modulator frequencies. Rather than waveforms, however, the ASSR assessment is based on a scalar measure of its phase-locking to the modulator (and its harmonics), usually expressed as a statistical quantity like an F-test.

While ASSR is faster for estimating thresholds than the standard serial ABR (Sininger et al., 2018), there are several practical aspects that have kept it from seeing widespread use in North America. ASSR does not provide waveforms, meaning it cannot be used to diagnose auditory neuropathy spectrum disorders (ANSD) or other retrocochlear lesions. The ASSR is also susceptible to artifacts spuriously detected as responses at high levels (Gorga et al., 2004), increasing the likelihood of misdiagnosed profound loss, a problem that becomes even more pronounced under bone conduction stimulus presentation (Swanepoel et al., 2008). Finally, the ASSR makes no use of audiologists’ expertise in interpreting waveforms, making many less inclined to use it.

The pABR provides full response waveforms rather than just a summary quantity, conferring several advantages over ASSR: 1) inference beyond the presence or absence of a response, such as individual response component amplitudes and latencies, 2) extended analysis windows and the separation of brainstem and cortical responses by their latencies, letting the clinician use the MLR to reinforce threshold estimates, and 3) minimal new training, as it draws on clinicians’ existing expertise in interpreting ABR waveforms.

### Next steps

#### The pABR as a platform for further innovation

The pABR paradigm is flexible, and can accommodate enhancements designed for serial ABR. For example, we have so far focused on toneburst stimuli due to their widespread use in North American clinics and well-established normative values for serial presentation. Band-limited chirp stimuli (Bell et al., 2002; Rodrigues et al., 2013) or other stimulus modifications, however, could easily be used in place of tonebursts, as long as the correction factors were also determined. Other frequency sets than the octaves from 500 to 8000 Hz used here could also be employed. For existing systems that use ER-3 earphones, the pABR upper range could be limited to 4000 Hz. Conversely, a recent study in rodents tested octaves from 1000 to 16,000 Hz, finding the pABR to be fast and accurate in that range as well (Suphinnapong et al., 2025).

Beyond simple parametric changes to stimuli, the pABR could also be coupled with other recent innovative techniques. Recent studies from two groups have developed waveform-based statistical methods for determining the presence or absence of a response (Shaheen et al., 2025; Undurraga, 2025). In concert with the pABR, these methods could enhance the information available to the clinician for determining thresholds, or could allow the pABR to run in an automated fashion similar to the ASSR. Other recent work has focused on optimizing the threshold search using Gaussian processes (Petersen and Shen, 2024; Chesnaye et al., 2025). While these methods were developed for serial ABR, adapting them to pABR, especially in concert with the aforementioned automatic response detection, could prove quite powerful.

In measuring all frequencies at a given level at once, the pABR provides opportunities for innovation that serial ABR does not. We recently developed a denoising tool that operates on the entire set of pABR waveforms, rather than considering each waveform separately (Maddox, 2025). Preliminary testing indicates a potentially substantial speedup over the threshold search paradigm used in this paper and could lead to new paradigms that would be especially useful in tele-medicine.

#### Translating the pABR

So far, work with pABR has been with air conduction stimuli, sensorineural hearing loss, and adult participants. Elevated thresholds measured with air conduction indicate a hearing loss, but not what type of hearing loss. Tympanometry can indicate conductive middle ear issues, but bone conduction ABR is necessary to determine the relative contributions of conductive and sensorineural loss. This distinction is important to make, as conductive losses may be temporary in infants due to, e.g., fluid in the ear. Even in chronic cases, the possible treatments and habilitation options differ for conductive versus sensorineural loss. Thus, bone conduction testing is important to evaluate with the pABR. Our preliminary work (Polonenko and Maddox, 2025) suggests that bone conduction is feasible with the pABR in adults with normal hearing, with similar waveforms for bone and air conduction stimuli when presented at the same dB nHL level for the respective transducer. That study also provides correction factors for bone conduction stimuli.

The most important work to be done is to test the pABR in infant participants with hearing loss. Given the proven accuracy of conventional ABR in infants and young children (Gorga et al., 2006; Stapells, 2011; McCreery et al., 2015) and the findings in adults from the present paper, there is reason to be optimistic about the pABR’s translation to infants. Still, changing paradigms in the clinic, which may also necessitate purchase of new equipment, is not undergone lightly. A study that provides compelling quantification of the pABR’s benefits, as well as age-appropriate norms and further refinement of the threshold search paradigm, will be a crucial step in the translation process.

## Data Availability

All data produced in the present study are available upon reasonable request to the authors.

## Declarations

### Author contribution statement

MP and RM designed the experiments, analyzed and interpreted the data, wrote and critically revised the paper. MP performed the experiments.

## Acknowledgements

This work was funded by the National Institutes of Health grant R01DC017962. Isabel Herb assisted data collection and Katherine Teece helped with recruitment. Eric Larson developed the interactive user interface. Portions of this article were presented at the following conferences: Molecules, Circuits, and Music: Communication Across the Auditory System Gordon Research Conference in Smithfield, Rhode Island in July 2024; 52nd Annual Scientific and Technology Conference of the American Auditory Society in Scottsdale, Arizona, USA on February 13, 2025; 48th Annual MidWinter Meeting of the Association for Research in Otolaryngology in Orlando, Florida, USA on February 26, 2025; Forum Acusticum EuroNoise in Malaga, Spain, on June 23, 2025; 2025 American Speech-Language-Hearing Association 2025 Convention in Washington, D.C, on November 20, 2025.

## References

Bell SL, Allen R, Lutman ME (2002) An investigation of the use of band-limited chirp stimuli to obtain the auditory brainstem response: Una investigatión sobre el uso de estímulos tipo “chirrido” (Chirp) de banda limitada para obtener respuestas auditivas del tallo cerebral. Int J Audiol 41:271–278.

Chesnaye MA, Simpson DM, Schlittenlacher J, Laugesen S, Bell SL (2025) Audiogram Estimation Performance Using Auditory Evoked Potentials and Gaussian Processes. Ear Hear 46:230.

Gorga MP, Johnson TA, Kaminski JR, Beauchaine KL, Garner CA, Neely ST (2006) Using a Combination of Click- and Tone Burst-Evoked Auditory Brain Stem Response Measurements to Estimate Pure-Tone Thresholds. Ear Hear 27:60–74.

Gorga MP, Neely ST, Hoover BM, Dierking DM, Beauchaine KL, Manning C (2004) Determining the Upper Limits of Stimulation for Auditory Steady-State Response Measurements. Ear Hear 25:302.

Hood LJ (1998) Clinical Applications of the Auditory Brainstem Response, 1st ed. San Diego: Singular Publishing.

Janssen RM, Usher L, Stapells DR (2010) The British Columbia’s Children’s Hospital Tone-Evoked Auditory Brainstem Response Protocol: How Long Do Infants Sleep and How Much Information Can Be Obtained in One Appointment? Ear Hear 31:722.

Kavanagh KT, Gould H, McCormick G, Franks R (1989) Comparison of the Identifiability of the Low Intensity ABR and MLR in the Mentally Handicapped Patient. Ear Hear 10:124.

Korczak P, Smart J, Delgado R, Strobel TM, Bradford C (2012) Auditory steady-state responses. J Am Acad Audiol 23:146–170.

Kothe C, Shirazi SY, Stenner T, Medine D, Boulay C, Grivich MI, Artoni F, Mullen T, Delorme A, Makeig S (2025) The Lab Streaming Layer for Synchronized Multimodal Recording. Imaging Neurosci 3:IMAG.a.136.

Luts H, Desloovere C, Wouters J (2006) Clinical application of dichotic multiple-stimulus auditory steady-state responses in high-risk newborns and young children. Audiol Neurootol 11:24–37.

Maddox RK (2025) Method and system for denoising sets of evoked potentials. Available at: https://patents.google.com/patent/WO2025072978A1/en?oq=WO+2025%2f072978+A1 [Accessed July 2, 2025].

McCreery RW, Kaminski J, Beauchaine K, Lenzen N, Simms K, Gorga MP (2015) The Impact of Degree of Hearing Loss on Auditory Brainstem Response Predictions of Behavioral Thresholds. Ear Hear 36:309.

Özdamar Ö, Kraus N (1983) Auditory Middle-Latency Responses in Humans. Audiology 22:34–49.

Petersen EA, Shen Y (2024) Multispecies initial numerical validation of an efficient algorithm prototype for auditory brainstem response hearing threshold estimation. J Acoust Soc Am 156:1674–1687.

Polonenko MJ, Maddox RK (2019) The Parallel Auditory Brainstem Response. Trends Hear 23:1–17.

Polonenko MJ, Maddox RK (2021) Optimizing Parameters for Using the Parallel Auditory Brainstem Response to Quickly Estimate Hearing Thresholds. Ear Hear 43:636–658.

Polonenko MJ, Maddox RK (2025) Bone conducted responses using the parallel auditory brainstem response (pABR) paradigm. :2025.10.07.680974 Available at: https://www.biorxiv.org/content/10.1101/2025.10.07.680974v1 [Accessed November 18, 2025].

Ramos N, Almeida MG, Lewis DR (2013) Correlation between frequency-specific auditory brainstem responses and behavioral hearing assessment in children with hearing loss. Rev CEFAC 15:796–802.

Rodrigues GRI, Ramos N, Lewis DR (2013) Comparing auditory brainstem responses (ABRs) to toneburst and narrow band CE-chirp® in young infants. Int J Pediatr Otorhinolaryngol 77:1555–1560.

Shaheen LA, Buran BN, Suthakar K, Koehler SD, Chung Y (2025) ABRpresto: An algorithm for automatic thresholding of the Auditory Brainstem Response using resampled cross-correlation across subaverages. Hear Res 462:109258.

Sininger YS, Hunter LL, Hayes D, Roush PA, Uhler KM (2018) Evaluation of Speed and Accuracy of Next-Generation Auditory Steady State Response and Auditory Brainstem Response Audiometry in Children With Normal Hearing and Hearing Loss: Ear Hear 39:1207–1223.

Stapells DR (2011) Frequency-specific ABR and ASSR threshold assessment in young infants. Phonak Com:409–448.

Stapells DR, Oates P (1997) Estimation of the Pure-Tone Audiogram by the Auditory Brainstem Response: A Review. Audiol Neurotol 2:257–280.

Stoll TJ, Maddox RK (2023) Enhanced Place Specificity of the Parallel Auditory Brainstem Response: A Modeling Study. Trends Hear 27:23312165231205719.

Stoll TJ, Maddox RK (2024) Enhanced Place Specificity of the Parallel Auditory Brainstem Response: An Electrophysiological Study. J Assoc Res Otolaryngol Available at: 10.1007/s10162-024-00959-w [Accessed October 16, 2024].

Suphinnapong P, Sabesan S, Lesica NA (2025) Estimating hearing thresholds in rodents using the parallel auditory brainstem response. Hear Res 461:109273.

Swanepoel DW, Ebrahim S, Friedland P, Swanepoel A, Pottas L (2008) Auditory steady-state responses to bone conduction stimuli in children with hearing loss. Int J Pediatr Otorhinolaryngol 72:1861–1871.

Undurraga JA (2025) Instrument for detecting auditory evoked neural responses. Available at: https://patents.google.com/patent/EP4509053A1/en?inventor=undurraga&assignee=interacoustics [Accessed November 18, 2025].

Vander Werff KR, Prieve BA, Georgantas LM (2009) Infant Air and Bone Conduction Tone Burst Auditory Brain Stem Responses for Classification of Hearing Loss and the Relationship to Behavioral Thresholds. Ear Hear 30:350.

